# The Influence of Contextual Factors on Maternal Healthcare Utilization in sub-Saharan Africa: A Scoping Review of Multilevel Models

**DOI:** 10.1101/2022.03.15.22272437

**Authors:** Simona Simona, Casildah Lumamba, Felicitas Moyo, Emily Ng’andu, Million Phiri

## Abstract

**Introduction:** Sub-Saharan Africa still bears the heaviest burden of maternal mortality among the regions of the world, with an estimated 201,000 (66%) women dying annually due to pregnancy and childbirth related complications. Utilisation of maternal healthcare services including antenatal care, skilled delivery and postnatal care contribute to a reduction of maternal and child mortality and morbidity. Factors influencing use of maternal healthcare occur at both the individual and contextual levels. The objective of this study was to systematically examine the evidence regarding the influence of contextual factors on uptake of maternal health care in sub-Saharan Africa.

**Materials and Methods:** The process of scoping review involved searching several electronic databases, identifying articles corresponding to the inclusion criteria and selecting them for extraction and analysis. Peer reviewed multilevel studies on maternal healthcare utilisation in sub-Saharan Africa published between 2007 and 2019 were selected. Two reviewers independently evaluated each study for inclusion and conflicts were resolved by consensus.

**Results:** We synthesised 34 studies that met the criteria of inclusion out of a total of 1,654 initial records. Most of the studies were single-country, cross-sectional in nature and involved two-level multilevel logistic regression models. The findings confirm the important role played by contextual factors in determining use of available maternal health care services in sub-Saharan Africa. The level of educational status, poverty, media exposure, autonomy, empowerment and access to health facilities within communities are some of the major drivers of use of maternal health services.

**Conclusions:** This review highlights the potential of addressing high-level factors in bolstering maternal health care utilisation in sub-Saharan Africa. Societies that prioritise the betterment of social conditions in communities and deal with the problematic gender norms will have a good chance of improving maternal health care utilisation and reducing maternal and child mortality. Better multilevel explanatory mechanisms that incorporates social theories are recommended in understanding use of maternal health care services in sub-Saharan Africa.

## Introduction

Sub-Saharan Africa (SSA) still bears the heaviest burden of maternal mortality, accounting for two-thirds of the world estimates [1]. Most maternal deaths occur during childbirth and the first one month of postpartum [2], which makes the pregnancy and childbirth periods crucial for the survival of mothers and new-born babies. SSA records disproportionately high maternal mortality mostly because of sub-optimal performance in not only maternal health service provision but also in the utilisation of those services during pregnancy and childbirth [3].

Evidence shows that maternal healthcare (MHC) services such as antenatal care, skilled delivery care and postnatal check-ups for mothers and babies are some of the most important drivers of maternal and child mortality in the world [3-5]. MHC helps provide health information that is necessary for healthy pregnancy outcomes [5]. It also ensures timely management and treatment of complications to minimise maternal deaths [4,6]. Despite the importance of MHC in ensuring the safety of both the mother and child, many women in SSA still face challenges using these services. For example, only about 52% of women benefit from skilled care during childbirth [7]. Studies have established many factors that prevent women from seeking health care during pregnancy and childbirth. These factors may be delineated at different levels, including the individual, heath systems and contextual levels. Several factors have been associated with maternal healthcare at the individual-level including maternal age [9] mother’s education [10], religion [11], family composition [12] wealth [4][10], information availability [13] place of residence [4][7], mothers’ occupation [12] health knowledge [14] and decision-making power [15] among others.

At health systems level, factors such as distance to the health facilities [17][18][19], perceived quality of healthcare [18] [20] cost [21], promptness of care [20], availability of medicine, equipment, and emergency care [19][20][22] are important in determining maternal healthcare in SSA. Contextual-level factors influencing maternal healthcare have included gender norms [23], poverty [24], cultural beliefs [7] government share of healthcare spending [25] and Gross National Income per capita [9].

Current research shows that variations in individual healthcare outcomes are significantly accounted for by contextual-level variables [13]. In fact, individual and health systems determinants of health and healthcare may be symptoms of much more ‘upstream’ contextual factors imbedded within local communities and broader social institutions. For example, the influence of women’s autonomy and decision-making authority on maternal healthcare have often been discussed within the framework of dominant masculinity ideologies or cultural beliefs in particular spaces [7]. The same may be true with inadequacies of the health systems, which may be a direct or indirect consequence of the failure of not the health systems per se but that of political and governance systems that oversee them.

There are many studies examining the relationship between contextual factors and maternal healthcare utilisation in continental SSA, but there are still inconsistencies relative to a) whether contextual factors influence maternal healthcare utilisation more than individual-level characteristics; b) What contextual factors are consistently associated with maternal healthcare utilisation and c) What theoretical pathways link contextual factors and maternal healthcare utilisation in SSA? These are the research gaps that this study intends to fill through a systematic review of multilevel models (MLMs).

MLMs have generally been regarded as ‘gold standards’ if the objective of the research inquiry is to examine the effects of high-level factors on individual-level outcome variables while controlling for other variables. This is because they are more suitable for determining the magnitude of variance in the outcome variable attributable to factors at different levels of operation [13]. Maternal healthcare utilization is underpinned by complex and interacting processes that may properly be explored by MLMs. Therefore, to examine the relative effect of contextual factors on maternal healthcare utilisation, we reviewed only multilevel studies that have included variables measured at two or more levels. This aspect distinguishes our study from previous reviews [26][7][12][27][28] which either were not entirely systematic, did not particularly focus on SSA, did not include all the indicators of maternal health care utilisation, or did not exclusively target studies using multilevel modelling.

Understanding the nature of contextual factors influencing individual women’s utilisation of MHC is critical for implementing of policy strategies aimed at bolstering the use of MHC services, especially in low-resource countries. Contextual factors provide a holistic way of looking at determinants of health and healthcare outcomes. Additionally, they provide a better platform for theoretical developments aimed at understanding the relationships between the broader social structure and individual level outcomes. This approach provides the potential to explain the disproportionately sub-optimal utilisation of maternal healthcare and high maternal mortality in SSA.

## Materials and Methods

As a matter of good practice, the protocol for this review was consistent with the Centre for Reviews and Dissemination (CRD) guidelines for conducting systematic reviews in healthcare [29]. The process involved searching the electronic literature, identifying articles corresponding to the inclusion criteria, and selecting them for extraction and analysis. As much as possible, this review adhered to the Preferred Reporting Items for Systematic Reviews and Meta-Analyses Protocols (PRISMA-P) 2015 statement checklist [30].

### Inclusion criteria

#### Types of studies

Although previous studies have used a variety of techniques to examine the influence of contextual factors on health and healthcare variables, this review searched only for studies utilising MLMs designs. MLMs designs are relatively new advances in quantitative research methods and best suited to study the relationship the effects of contextual conditions on individual-level health outcomes because they allow for a simultaneous examination of the relative impact of individual level and group level variables on the outcome variable [31][32]. They also accommodate an analysis of observations that are correlated or clustered along spatial, non-spatial, or temporal dimensions [33]. The application of MLMs in such data structures helps to model the extent of correlation in the outcome attributable to contextual factors while controlling for individual-level variables [13]. These properties of MLMs enabled us to determine contextual factors associated with maternal healthcare and how the impact of these factors differs from individual-level variables. Studies that used any type of MLM techniques and focused on a single or multiple sub-Saharan African countries were eligible for selection.

#### Publication status

Studies published in refereed journals between 2007 and 2019 were included. It was assumed that this period would capture adequate empirical studies applying MLMs in sub-Saharan Africa. Papers and materials published in non-refereed journals, book reviews, unpublished working papers, government reports, qualitative studies, dissertations, published comments, expert opinions, media briefings, editorials and book reviews were excluded. Studies that use MLMs but did not measure any higher-level variable (s) were not included in this review. The review included only studies published in the English language.

#### Participants and setting

The review included studies that were implemented in any sub-Saharan Africa country, as defined by the World Bank Group [34], focusing on maternal healthcare utilisation. This review excluded studies which targeted low and income-countries that included other sub-Saharan citizens in the diaspora.

#### Outcomes

Studies reporting maternal healthcare utilisation as the outcome variable of concern were included in the review. The indicators of maternal healthcare utilisation were restricted to contraceptive use, antenatal care, skilled/facility delivery care and postnatal care.

#### Determinants

Contextual-level determinants of maternal healthcare utilisation in MLM studies were considered suitable, and these included factors clustered at the household, community, district, regional or national levels. Characteristics such as community socioeconomic status, social norms, social cohesion and national level economic development status, poverty levels and health expenditure were among eligible contextual factors.

### Information sources

To avoid bias and to reflect the multidisciplinary nature of the subject of this review, literature searches were performed across a variety of databases including Science Direct, ProQuest, Google scholar, MEDLINE, EMBASE, PubMed, WHO Library Database (WHOLIS), African Health line, Wiley Online Library, (AIM), Web of Science, and Scopus

### Literature search strategy

This review employed four sets of terms representing the outcome indicator variables, the contextual indicator variables, MLM filters and the filter for sub-Saharan African countries in the search strategy. Table 1 below reports an example of the search strategy adopted for this review. The search filter for sub-Saharan African countries was adapted from work by Pienaar, et al [35] who developed it for clinical studies conducted in an African environment. The filter comprised sub-Saharan African country names and truncated terms such as “Southern Africa”. North African countries and language filters were removed from the original list accordingly.

**Table 1.** Example of search strategy.

|  |  |
| --- | --- |
| Maternal healthcare indicator terms | “Maternal healthcare” OR “access to healthcare” OR “availability of healthcare” OR “utilisation of healthcare” OR “health service coverage” OR “contraceptive use” OR “skilled birth attendant” OR “skilled delivery care” OR “facility based delivery” OR “antenatal care” |
| Contextual factors | “Structural factors” OR “social determinants” OR “contextual factors” OR “determinants” OR “neighbourhood status” OR “community factors” OR “structural violence” OR “social structure” OR “social norms” OR “sociocultural factors” OR |
|  | "economic development" OR "governance" "gender inequality" OR "predictors" OR "health systems" |
| Multilevel models filters | "Multilevel" OR " multilevel models" OR "multilevel modelling" OR "MLM" OR " hierarchical models" OR "hierarchical linear models" OR "HLM" "Variance component" OR "Intraclass correlation" OR "ICC" OR "random effects" OR "mixed models" OR "mixed effects" |
| Filter for sub-Saharan African countries | ("Africa" OR Angola OR Benin OR Botswana OR "Burkina Faso" OR Burundi OR Cameroon OR "Canary Islands" OR "Cape Verde" OR "Central African Republic" OR Chad OR Comoros OR Congo OR "Democratic Republic of Congo" OR Djibouti OR "Equatorial Guinea" OR Eritrea OR Ethiopia OR Gabon OR Gambia OR Ghana OR Guinea OR "Guinea Bissau" OR "Ivory Coast" OR "Cote d'Ivoire" OR Kenya OR Lesotho OR Liberia OR Madagascar OR Malawi OR Mali OR Mauritania OR Mauritius OR Mozambique OR Namibia OR Niger OR Nigeria OR Principe OR Reunion OR Rwanda OR "Sao Tome" OR Senegal OR Seychelles OR "Sierra Leone" OR Somalia OR "South Africa" OR Sudan OR Swaziland OR Tanzania OR Togo OR Tunisia OR Uganda OR "Western Sahara" OR Zaire OR Zambia OR Zimbabwe OR "Central Africa" OR "Central African" OR "West Africa" OR "West African" OR "Western Africa" OR "Western African" OR "East Africa" OR "East African" OR "Eastern Africa" OR "Eastern African" OR "North Africa" OR "Southern Africa" OR "sub Saharan Africa") |

### Data extraction and management

A search was conducted in databases using the strategy above to identify potential records for the review. All extracted entries were screened by examining titles and abstracts and all relevant records with the potential of meeting the criteria for inclusion were exported to EndNote X7 for data management. Full texts were extracted using the EndNote platform and those, which could not be found, were searched manually through the university library and other sources. Full texts were assessed by two independent researchers against the inclusion criteria and disagreements, where there were any, were resolved by consensus. Studies excluded from the review at this stage, along with reasons for exclusion, were recorded and retained by the researcher. The search also included hand searching reference lists of relevant articles to identify other articles of value.

Records fitting the inclusion criteria were then exported to excel to record the detailed general publication information (author(s), year of publication and type of journal), characteristics of studies (design, population, sample size and procedure, country, explanatory, and outcome variables of concern) and summary results (whether contextual factors significantly influenced maternal healthcare utilisation and recording the intra-class correlation ICC or variance partition coefficient VPC). The search was conducted on the 16^th^ of April 2018, and it was updated on the 14^th^ of June 2021.

### Risks of bias and quality assessment

Poor quality assessment of studies has a considerable negative impact on the results of systematic reviews. As such, this review appraised all studies in accordance with the Quality Assessment Tool for Quantitative Studies developed by the Effective Public Health Practice Project [36]. Accordingly, the assessment considered selection, the appropriateness of study designs, whether there were stated confounders, whether there were variables which had been controlled for, whether blinding was applied, reliability and validity of data collection tools, the appropriateness of the units of analysis and the type of variables included. The way missing data were resolved was also part of the assessment procedures.

Just like the process of data extraction, two reviewers were involved in the quality assessment in the final processes of this review and in cases of disagreement, a third reviewer was enlisted to reach consensus. In cases of missing data, attempts were made to contact the primary authors of articles concerned. This was done in situations where, for example, study designs, explanatory or outcome variables were unclear or missing. When there was no success in obtaining the missing data, data were reported accordingly, and the implication thereof explained in the discussion section of the review.

### Data analysis and presentation

Heterogeneity in the study population, selected countries, outcomes, and the nature of contextual variables was observed. As such, meta-analysis was not appropriate for this synthesis. Instead, narrative scoping review was employed as the method of reporting findings. A table was created with six columns and used to report the name of first author and reference in column 1. Contextual or structural factors that were examined after controlling for individual-level variables in column 2. Individual factors from adjusted models were reported in column 3. Column 4 reported the Intra-class correlation (ICC), which is an inferential statistic measuring variability within contextual factors influencing maternal healthcare utilisation. It is used here to determine the degree of variation in use of maternal health care attributed to contextual-level factors. Column 5 reports the outcome variable investigated in the research and the last column shows the direction of the relationship between significant contextual/structural factors and indicators of maternal healthcare under consideration.

## Results

### Search results

The electronic data search process from the research platforms above yielded 1,654 potentially relevant records. Twenty-seven additional records were identified from the reference lists and citation checks of included studies. After searching for duplicates, 996 were discarded and 658 records were screened using titles and abstracts. Following this stage, 591 irrelevant manuscripts were excluded from the review and the remaining 67 records were fully assessed with respect to the illegibility criteria and among these, thirty-nine were selected. The main reasons for exclusion included studies not focusing wholly on sub-Saharan Africa, not using multilevel modelling techniques, using primarily qualitative methods, not being published in a peer-reviewed outlet, and having an outcome variable other than one of the indicators of maternal healthcare. After quality assessment for confounding and applying the measurement of contextual variables requirement in MLM studies, five were excluded and the remaining 34 records were eligible for inclusion in the study. The figure below is a flow diagram, which reports the process from article identification to inclusion.

### Study characteristics

Apart from a few entries [37][38][39][40][41], almost all reviewed studies used the Demographic and Health Surveys (DHS) data from a standard survey that is implemented in several developing countries. McTavish and others [40] is the only study using the World Health Survey (WHS) and controls for the national level socioeconomic characteristics when investigating the relationship between national female literacy and maternal healthcare use. Table 2 reports the distribution of selected studies across nine sub-Saharan African countries, including some cross-national studies, which used a combination of SSA countries as a unit of analysis.

**Table 2.** Distribution of studies by country.

| Country | Count | Percent |
| --- | --- | --- |
| Ethiopia | 7 | 20.6 |
| Nigeria | 6 | 17.6 |
| Multi-country | 5 | 14.7 |
| Tanzania | 4 | 11.8 |
| Ghana | 3 | 8.8 |
| Mali | 2 | 5.9 |
| Zambia | 2 | 5.9 |
| Mozambique | 2 | 5.9 |
| Kenya | 1 | 2.9 |
| Zimbabwe | 1 | 2.9 |
| South Africa | 1 | 2.9 |
| **Total** | **34** | **100.0** |

Sample sizes for included studies have a wide range because of heterogeneity in these studies. Some studies, for example, only focus on a relatively small rural population in one country [38][39][40][42][43] while some others pool data in a cross-national design, making the total sample size relatively large [13][23][41][44]. However, all the selected studies have sample sizes adequate to represent populations under investigation.

Reviewed studies are mainly cross-sectional and utilized multilevel logistic regression analysis with a few making use of multinomial and structural equation regression analysis. This is plausible because of’ lack of robust data infrastructure in SSA which prohibits the use of more comprehensive and advanced analytical techniques and could be the reason why studies using such methods as longitudinal multilevel analysis are missing.

Table 3 reports the results of the systematic review in terms of the study references, factors influencing different indicators of maternal healthcare, including both contextual and individual-level factors controlled for. The ICC and the direction of the relationship between contextual-level factors and maternal healthcare are also reported. There is observed heterogeneity in the constitution of contextual factors in included studies. However, in most studies, these factors are constituted by aggregating individual-level data to represent community characteristics often defined as clusters or PSUs in the case of the demographic and health surveys (DHS) data [23][24][44][45][46][47]. Studies by Balew et al, [47] and Ngome and Odimegwu [48] define contextual factors as structural determinants and social contexts respectively and focus exclusively on their influence on maternal healthcare. Most of the other studies seek to investigate the effects of both individual and community-level factors and emphasize the importance of considering factors operating at both levels in policy strategies aimed at bolstering maternal healthcare use.

**Table 3.** Summary of studies included in the systematic review.

| Study (Year) | Main contextual factors for adjusted models | Individual level factors for adjusted models | ICC | Outcome | Direction of contextual influence |
| --- | --- | --- | --- | --- | --- |
| Adjiwanou & LeGrand 2014[23] | Gender norms regarding violence against women | Women's education, maternal age, birth order, spouse education, wealth index, employment | [0.40]<sup>a</sup> | Skilled birth attendance, 4+ ANC visits and first trimester ANC | Women who live in areas where gender norms are pronounced are less likely to use skilled birth attendants and timely antenatal care in Ghana, Tanzania, and Uganda |
| Chama-Chiliba & Koch 2015 [45] | Area of residence, quality of health care<sup>b</sup> | Employment status, partner education, household childcare burden, quality of ANC received | [0.11] | Utilization of antenatal care | High quality healthcare is associated with insufficient ANC visits. In the rural areas this has more explanatory power than in urban areas |
| Masters et al 2013 [46] | Travel time to health facilities | Maternal education, age, household wealth, female autonomy, partner education, parity | [nm] | In-facility delivery and antenatal care visits | An increase in travel-time reduces the odds of in-facility delivery and antenatal care visits |
| Balew et al 2015 [47] | Religion, place of residence and media exposure | Maternal education, household wealth | [0.40] | Family planning services | Less media exposure, living in rural areas belonging to Islam and other religions lowers the probability of using family planning services at the community level |
| Girmaye & Berhan 2016 [43] | Distance to the nearest health provider | Skilled personnel preferred, awareness of providers, media exposure | [0.60] | Skilled antenatal care services | The further the distance to the nearest provider the less the likelihood to use skilled antenatal care |
| Gage et al 2016 [49] | Health system, local government areas | Maternal education, place of residence, household wealth | [0.33] | Health facility delivery | Women using health systems and in local government areas with supportive quality of MHC have higher likelihood of having HF delivery |
| Gage 2007 [50] | Area effects: Access to facility, availability of health facility, social environment, poverty, region | Household type, childcare burden, mother characteristics, birth order | [0.42]<sup>c</sup> | Receipt of prenatal care, prenatal visits, skilled delivery, institutional delivery | Living closer to other women who used MHC services, having transport, and staying in a place with more educated women increases the odds of utilising maternal health services. |
| Afulani 2015 [51] | Socioeconomic status, Place of residence | Urban living, higher education, higher wealth, 4+ANC visits, ANC with a doctor, ANC in government hospital, used contraceptives | [0.27]<br>[0.15] | Quality of antenatal care | Women who live in areas with higher socioeconomic status have better chance of utilising higher quality antenatal care |
| Elfstrom & Stephenson 2012 [44] | Fertility knowledge, gender norms and inequalities, health knowledge | Age, parity, woman's education, household wealth | [nm] | Contraceptive use in Africa | Women living in community with higher fertility knowledge, unproblematic gender norms and have health knowledge are more likely to use modern contraceptives |
| Kaggwa et al 2008 [52] | Proportions of women in the community who: are exposed to FP messages, desire small family size, have access to piped water | Age, education, religion, place of residence, ethnicity, socioeconomic status, approval of family planning | [0.23] | Contraceptive use | Exposure to FP messages and number of births per woman were associated with contraceptive use. After adjusting for individual level factors, none of contextual level factors were associated with contraceptive use in Mali |
| Ndao-Brumblay et al 2012 [40] | Village characteristics: Distance to health facility, village poverty, road network quality | Age, marital status, household wealth index, optimal antenatal visits, parity, education, health insurance | [0.36] | Institutional or health facility delivery | Contextual level factors were not significant in explaining variations in the use of health facility for delivery |
| Dias & de Oliveira 2015 [53] | Average community wealth and Social economic position | Female occupation, age, distance to health facility, marital status, religion, education, wealth | [0.09] | Women's use of modern Contraceptives | Women's socioeconomic position and average community wealth are positively associated with use of use of modern contraceptives |
| Ononokpono 2015 [54] | Region of residence, community education, poverty, hospital delivery, media exposure | Maternal age, education, women's autonomy, household wealth, occupation, religion, ethnic origin, place of residence | [0.66] | Skilled antenatal care | Region, community hospital delivery, poverty and media exposure have strong independent associations with skilled antenatal care |
| Ononokpono et al 2014 [55] | Community education, community health facility delivery and ethnic diversity | Maternal age, educational attainment, religion, ethnic origin, occupation, household wealth, household size | [0.42] | Postnatal care | Living in communities with high proportion of educated women, those who had facility delivery, increases the likelihood of postnatal care |
| Ononokpono et al 2013 [56] | Community health facility delivery, community poverty and education | Women occupation, household wealth, women autonomy and region of residence | [0.47] | ANC visits | Living in community with high facility delivery increases the chance of 4+ ANC visits while community poverty lowers the likelihood of antenatal care visits |
| Ononokpono & Odimegwu 2014 [57] | Place or residence, region of residence, community women's education, ethnicity diversity | Age, educational attainment, religion, ethnic origin, religion, education, household index, parity, women autonomy | [0.69] | Health facility delivery care | Women from rural areas, those from north east, north west, south south and south west are more likely to have facility delivery. Furthermore, communities with more educated people increase the odds of facility delivery |
| Worku et al 2013 [39] | Community: Source of income, distance to health centre, payment during delivery, obstetric guideline, self-sustained, duty service | Birth order, educational status, wealth, preference for skilled provider, awareness of risk of pregnancy, previous experience of ANC | [0.31]<sup>d</sup> | Antenatal, delivery and postnatal care | Payment requirement, distance to the facility are significant barriers to skilled delivery. Health facility performance was significantly associated with use of skilled maternal care. |
| Debelew et al 2014 [58] | Place of residence, distance from health centre and hospital | Age, Educational status, household wealth, religion, ethnicity, occupational status | [0.55] | Birth preparedness and complication readiness | Urban residence and shorter distance to the hospital increase the likelihood of birth preparedness and complication readiness |
| Babalola & Fatusi 2009 [59] | Place of residence, media exposure, small family norm | Women's Education, socioeconomic status, maternal age, ideal family size | [0.40]<sup>e</sup> | Use of antenatal care, medical personnel delivery, and postnatal services in Nigeria | Urban residence and community media saturation are strong predictors of use of maternal healthcare services |
| Mohan et al 2015 [60] | Community: Trust, education, poverty, family planning, distance, and utilization of health facility | Education, age, birth order, antenatal visits, history of complications, place of delivery, counselling received | [0.20] | Postnatal care use at health facility | High community levels of postpartum family planning usage and high level of trust in health system increase the likelihood of using postnatal care services |
| Stephenson et al 2008 [61] | Distance to health facility, male/female education ratio, mean age at marriage and reporting physical violence, PSU household size | Place of residence, age, place of residence, parity, education, marital status, race, employment, household size, partner approval, | [0.30] | Modern contraceptive methods | Women living further away from health facilities were more likely to use contraceptive methods. Living in communities with more educated men than women reduce the odds of contraceptive use.<br>Living in communities with higher age at marriage and with more women reporting partner violence increased contraceptive use |
| Cau 2015 [62] | Community: Exposure to FP messages, women empowerment, women autonomy, fertility desire, place of residence | Parity, age, professional status, education, desired family size, marital status, marriage type, exposure to FP messages | [0.06] | Contraceptive use | Women's empowerment and fertility desires in the community significantly increases the odds of using modern contraceptive methods. |
| Yebyo et al 2015 [24] | Place of residence, women empowerment, | Household wealth, educational attainment, partner education, | [0.75] | Home delivery | Rural, agrarian, and poor communities as well as communities with longer distance |
|  | education levels, media exposure, ANC visits, distance to health facility | media exposure and perceived problems with transportation |  |  | from health facilities and lower ANC utilization had higher odds of home delivery. |
| Achia & Mageto 2015 [63] | Place of residence, region of residence, socio-economic status, community population density, facility delivery index, community diversity | Age, maternal education, religion, marital status, ethnicity, partner education, socio-economic status, decision-making authority, media exposure, insurance coverage, employment status | [0.25] | Adequate use of antenatal care, skilled delivery care | Greater community ethnic diversity, higher community socio-economic status, community health facility delivery was associated with adequate use of maternal health care |
| Kruk et al 2015 [42] | Village farms cash crops, distance to hospital, number of shops and water pumps, population size | Education, healthcare utilisation, media exposure, age, marital status, religion, household assets, self-rated health, occupation | [0.27] | Home delivery | Farming of cash crop in a village reduces the probability of home delivery. Longer distances from the hospital increases odds of home delivery |
| Kruk et al 2010 [37] | Access to health facility, community perception of the quality of local health system | Age, education, wealth status, insurance, use of healthcare, parity, health beliefs, perceptions of a health system, | [0.34] | Facility delivery | Positive perceptions of health facilities and skills of health professionals in the villages, inversely negative perception of traditional birth attend skills increases the likelihood of facility delivery |
| McTavish 2010 [41] | National female literacy | Individual age, education, urbanicity and household income, marital/cohabitation status | [0.20] | Use maternal health care | Mothers residing in countries with high levels of female literacy were more likely to use maternal health care |
| Jacobs et al 2017 [38] | Type of residence, ratio of facility to population | Age, school attendance, marital status, literacy, | [0.22]<sup>f</sup> | Antenatal care, skilled delivery, postnatal care | Women who live in centralized rural areas were more likely to use SBA than those living in remote rural areas. The other community-level variable was not significant in all indicators |
| Mezmur et al 2017 [64] | Community: education, poverty, level of fertility, level of contraceptive prevalence, level of health service utilization and place of residence | Age group, birth order, ethnic origin, religion, maternal education, media access, occupation, and wealth index | [0.61]<sup>g</sup> | Skilled birth attendance (SBA) | place of residence, community level of female education and fertility significantly influence the uptake of skilled delivery care |
| Adu et al 2018 [65] | Place of residence, Community education, community wealth, distance to health facility, having insurance | Age, ethnicity, religion, marital status, parity | [0.17] | Antenatal, delivery and postnatal care | Women in communities with higher education were more likely to access antenatal and skilled delivery care. Women in wealthy communities were more likely to access postnatal care |
| Ngome & Odimegwu 2014 [48] | Provincial level: access to healthcare, education, number of children per women, ANC attendance | Women's autonomy, age, marital status, education, media access, parity, place of residence, household wealth, parity | [0.18] | use of modern contraceptives | Lack of access to healthcare, more children per women and having higher education are associated with less use of modern contraceptives in a province |
| Yaya et al 2018 [66] | Neighbourhood SES, Human Development Index (HDI) | Women empowerment, age, wealth, partner education, decision-making power | [0.39]<br>[0.21] | Contraceptive use | Women in less disadvantaged neighbourhoods were significantly (57%) less likely to have used contraceptives compared to those in least disadvantaged spaces. Women in moderate HDI countries are more likely to use contraceptives than those from low HDI countries |
| Tegegne et al 2019 [67] | Residence, antenatal care readiness, service availability, distance to facility | Age, education, occupation, husband education, household head, family size, wealth, parity, autonomy, | [0.25] | Use of antenatal care, | Living in rural areas with longer distance to health facility reduces the likelihood of using antenatal care in |
| Woldegiorgis et al 2019 [68] | Urban population, literacy rate, ratio of health professionals to population, income grouping | Age, marital status, partner education, literacy, number of ANC, birth spacing, residence, wealth, and perceived need | nm | Antenatal care (ANC) visits and skilled birth attendance (SBA) | Women in middle income countries were more likely to have more ANC visits and access SBA compared to those from low-income countries. Countries with higher literacy rate have decreased association with ANC visits and SBA |

<sup>a</sup>The highest reported ICC among the four countries studied
<sup>b</sup>Meaning drug availability and density of health facilities
<sup>c</sup>For institutional delivery, the highest among the four outcomes (others are: receipt of prenatal care: 0.09, prenatal visits: 0.17, skilled delivery: 0.39)
<sup>d</sup>For antenatal care, highest among the maternal health indicators studied (Others are delivery care: 0.12 and postnatal care: 0.25)
<sup>e</sup>ICC for skilled delivery (other outcome indicators: antenatal care: 0.37; postnatal care: 0.34)
<sup>f</sup>ICC for skilled birth attendance (other out indicators: antenatal care: 0.09 and postnatal care: 0.22)
<sup>nm</sup>Not measured

Elfstrom and Stephenson [44] and Masters [46] are examples of studies using a three-level modelling technique in contrast with most studies using two-level models. Both studies assume that individual actors are nested within households which, are in turn nested within communities. They are interested in the influence of contextual factors on contraceptive use at the community level of analysis and they studied demographics and fertility norms, gender norms and health knowledge. These studies included household and individual-level factors, which were used as control variables.

Outcome variables in included studies are equally varied, covering the broader spectrum of maternal healthcare continuum of contraceptive use [44,48,52,53,61,62,66], antenatal care [43,45,51,54,56,67] skilled or facility delivery [24,37,40,42,47,57,64], and postnatal care [55,60]. Some studies included a combination of two maternal healthcare indicators as outcome variables of antenatal care and facility/skilled delivery [23,46,50,63,68] or three indicators of antenatal care, skilled delivery and postnatal care [38,39,59,63,65].

### Contextual factors influencing maternal healthcare utilization

Table 3 also reports several contextual factors that were found to be associated with maternal healthcare after controlling for individual level characteristics. These have been grouped into four categories, including health systems, relational factors, and community socioeconomic position and cultural. Health systems variables are those that relate to the location, capacity, quality and The most common health systems level factors at the community level include distance or access to health facilities [24][37][40][42][43] [46][48][50][58][60][61][65][67] and quality of healthcare offered [37][38[45] [64][67][68]. Quality of health care is broadly defined to include the ratio of healthcare professionals to population, perceptions of the quality of local health facilities, care readiness and service availability.

The overarching nature of the relationship between health systems and maternal healthcare is such that women who live in communities that are proximal to well-performing and high-quality health systems have higher odds of utilizing maternal healthcare services. It was unusual that a few studies either found a negative association between distance and maternal health care [61] or the relationship was non-significant [37][61]. Higher levels of facility delivery in these cases were explained by the quality of healthcare, which seems to supersede distance as an explanatory factor for facility delivery. Other health care-related factors that were found to increase the odds of utilizing maternal health care system included trust in the health care system [60] and the lack of payment requirements [37].

A host of socioeconomic factors were found to be associated with maternal healthcare utilization at the contextual level, and these included community education levels[24][54][55][56][57][60][65], community wealth levels or poverty [53][57][60] [65], population density [45][63][65], community media exposure or exposure to family planning messages [24][47][52][54][64], place of residence [24][45][47][52][54][57] [58][59][62][63][65][67], and gender norms [23][44]. Most of these factors have a consisted and predictable relationship with maternal healthcare. For example, it is well-known that women who live in communities or countries with higher levels of education or economic status have increased odds of utilizing maternal healthcare.

On the other hand, it is also established that women who live in rural areas have a lower propensity of using maternal healthcare services. The same applies to community media exposure, which has mostly been found to have a positive influence on maternal health care. Problematic gender norms such as tolerance towards violence hinder women from using maternal healthcare.

Relational factors are also found to be important predictors of maternal healthcare utilization at the contextual level. These factors mainly result from everyday relationships or interactions women have with men. These relations may significantly influence women’s decisions to either use maternal healthcare or not. Community women empowerment [24][62], women autonomy [62], PSU household size [61], number of children per PSU [48] and small family norms [52][59] are among the most consistent predictors of maternal healthcare utilization.

The findings show that in sub-Saharan Africa, residence in communities where most women are empowered is advantageous in terms of use and access to maternal healthcare services. It is also evident that women who live in areas where their individual freedoms are inhibited have less likelihood of using maternal healthcare. The results also indicate that contextual small family norms are positively associated with contraceptive use.

To measure the independent variations in maternal healthcare attributable to contextual factors, we observed the value of the Intra-class Correlation Coefficients (ICC) for the contextual level factors in MLMs of reviewed studies. Included studies have not been consistent in reporting ICC measurements, which indicate the relative importance of contextual factors to individual-level factors in explaining variations in the use of maternal healthcare services. We extracted the ICC values from null models (i.e. before individual and contextual level variables are included) in all included studies. Where the ICC was not reported at all, it was computed using available variance partition information.

The ICC results reported in Figure 2 show that they are averaging 35%, which indicates that relative to individual-level factors, contextual level factors contribute less to maternal healthcare utilization but have substantial independent influence. There are a few studies which are above the ICC of 50% [24][43][54][57][58][64] and this show more impact attributed to contextual factors than individual-level characteristics. These studies are addressing antenatal care and skilled facility delivery using several contextual factors, including place of residence, women empowerment, community education, level of fertility, distance to health facility, poverty, and community media exposure, among others.

**Figure 1.**
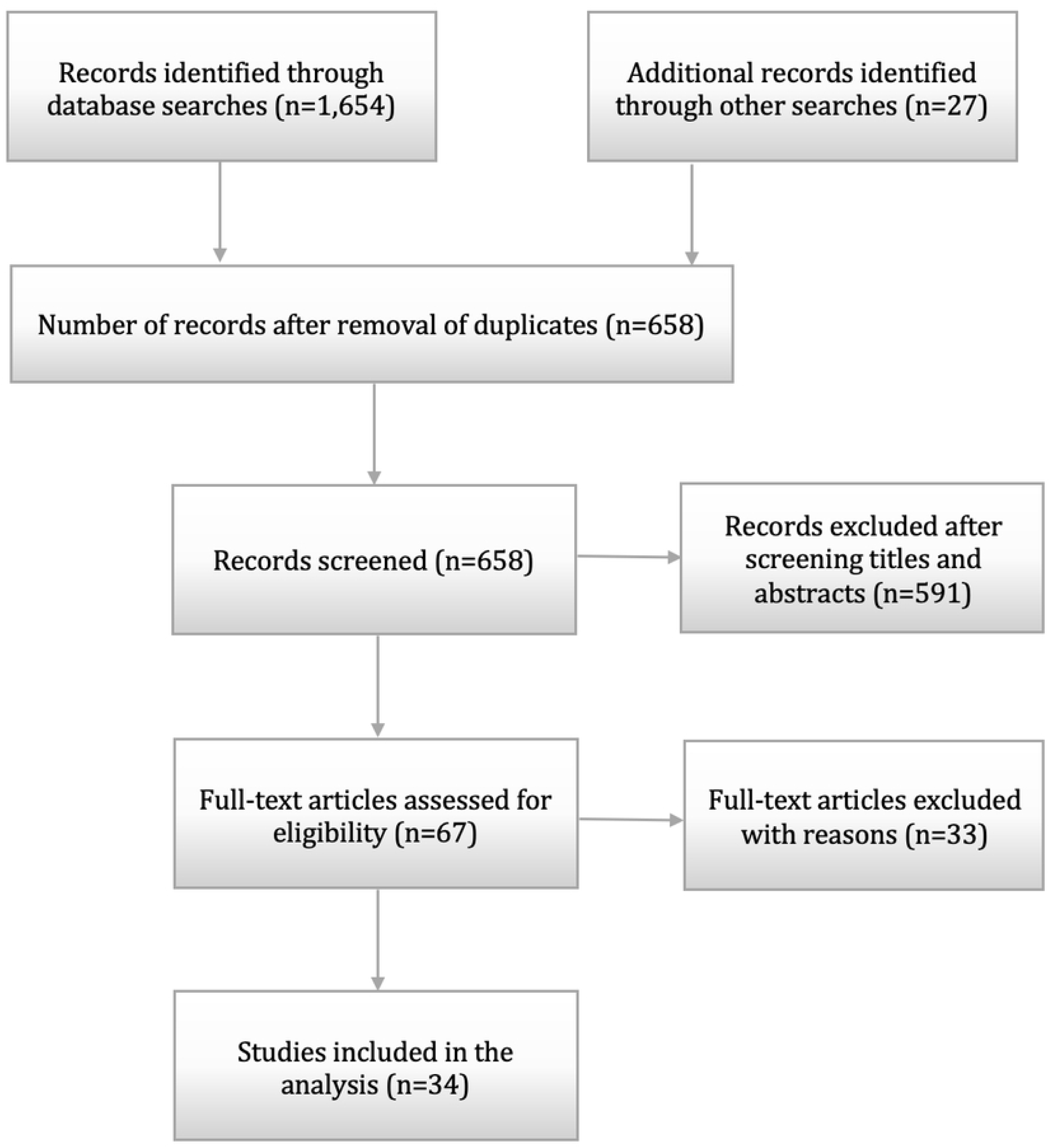
Flow of information through different phases of systematic review. Source: Moher et al, [30].

**Figure 2.**
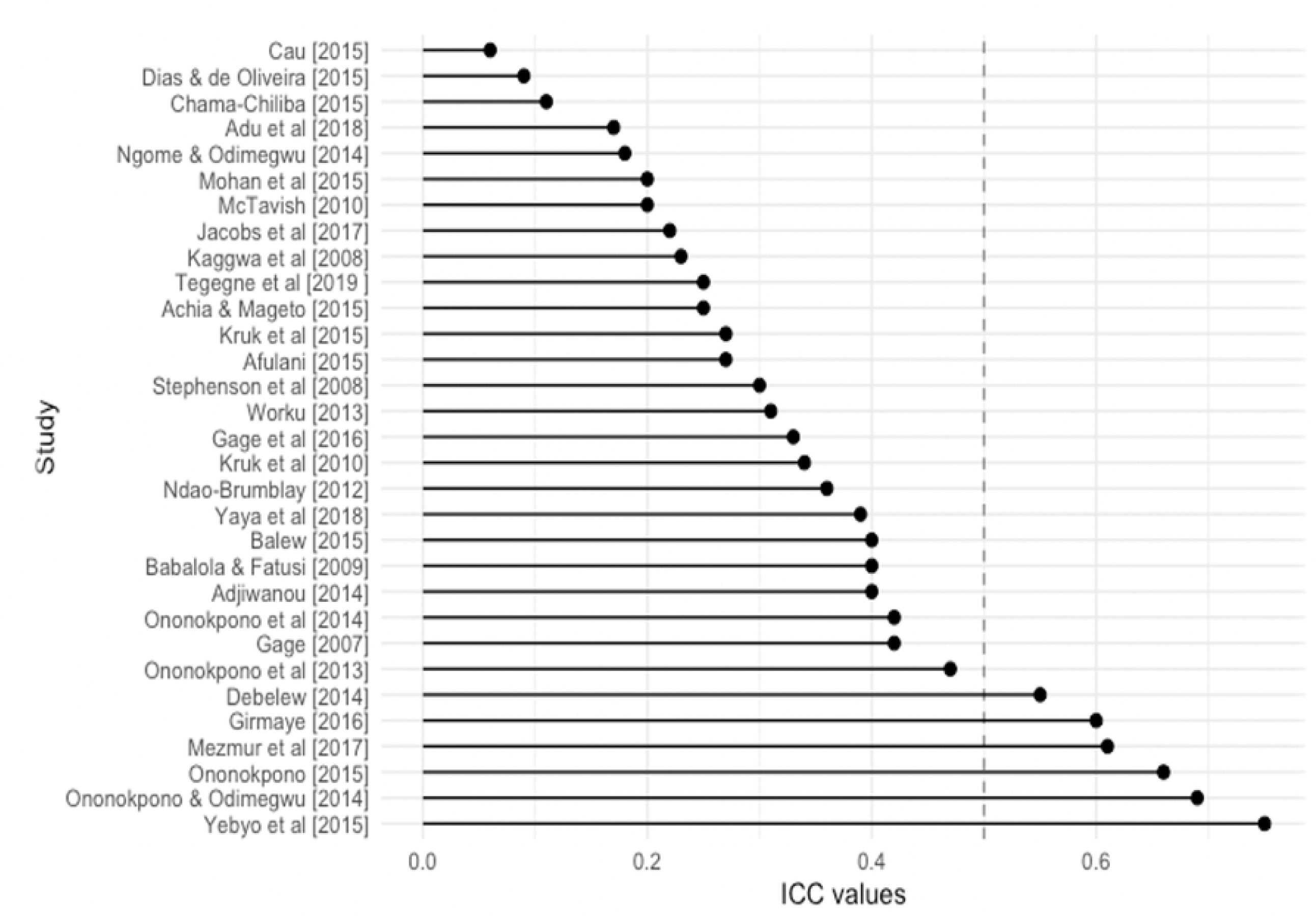
ICC values by study.

### Theoretical pathways linking contextual factors and maternal healthcare utilization

We sought to identify theoretical underpinnings behind observed relationships between contextual factors and maternal healthcare utilisation in SSA. Reviewed studies have mostly been sound in identifying the joint effects of individual and contextual level factors associated with maternal health care [24][38][39][54][57][64]. However, there are still challenges in articulating the underlying mechanisms behind these relationships. What, for example, are the social mechanisms making women living in underprivileged, less educated, or rural communities have inadequate maternal healthcare utilization? There are only a few studies that offer plausible explanations to unpack such findings. Adu and others [65] for example, attempt to explain the influence of educated communities in maternal health care utilization by illuminating the potential shift in community power structures that may result in more autonomy for women and subsequently better use of health services.

Kruk and others have equally attempted to do so by demonstrating the role played by contextual social interactions as a mechanism through which women make decisions to utilize maternal health care services [37]. They argue that, as women interact within communities, they tend to share information about the quality of health care systems and these collective perceptions shape women’s decision-making processes about use of health services. Furthermore, a few other studies that have used contextual social norms in explaining variations in health service utilization [23][42] help to illuminate the role of the social structure and how it is distinctive of individual characteristics in influencing health behaviour. This is an insightful approach as it gives a broader picture through which the observed relationships are shaped by prevailing social and cultural factors in a particular society.

Most reviewed papers do not offer strong theoretical frameworks on the influence of contextual factors on maternal health care utilization. This may be partly because of inability to articulate the presence of emergent contextual properties that are distinctive from individual-level constructs [69]. Lack of clear-cut distinction between contextual and individual-level constructs can result in atomistic fallacy, whereby conclusions about contextual-level variables are made from individual-level data [70]. There are many instances among reviewed studies where researchers are discussing contextual effects with individual-level data. This is problematic, and it poses a challenge in grounding such studies in theoretical constructs.

## Discussion

This scoping review used multilevel models in sub-Saharan Africa to study the influence of contextual factors on maternal healthcare utilisation. Our synthesis involved 34 studies that met the criteria and controlled for some individual-level factors. Previous reviews focused on different indicators of maternal healthcare, did not particularly look at SSA or did not specifically target multilevel models [7][12][26][27]. Selected studies in this review reflected substantial heterogeneity in terms of contextual factors and outcome variables examined. Contextual factors are defined as structural-level factors constituted at the community or country levels and are mainly studied through multilevel modelling techniques. Contextual factors found to be commonly associated with maternal healthcare relate to the operations of health systems, socioeconomic positions of women, and the nature of relationships between women and men.

The health system factors show that women who live in areas that are proximal to health facilities, provide good quality healthcare, have full antenatal care coverage and, where more people trust in the health system, have a higher propensity to use maternal healthcare services. These findings imply that access and performance of health systems in SSA are important in facilitating optimal use of health services among pregnant women, mothers, and newly born babies. However, poor performance of health systems may be due to broader structural factors, such as the political and governance systems prevailing in specific countries of SSA. This is plausible because healthcare delivery capacity is considerably less in developing countries compared to developed countries. This may result from low health expenditure per capita, which in 2015, was approximately $37 in low-income countries compared to around $518 and $5,251 in upper middle-income countries and higher income countries respectively [16].

Consequently, the health professional to population ratio is also less impressive as it stands at 0.2 for physicians and 1.2 for nurses and midwives in SSA countries, while the corresponding figures for the developed countries are 2.9 and 8.6 per thousand population [16]. These phenomena may account for the suboptimal performance of health systems in SSA.

Urban residence or living in areas with more educated and wealthy women could enhance the propensity of using maternal health care [43][24][56][54]. Urban residence and high socioeconomic status are usually associated with good health outcomes because, on one hand, better amenities are usually found in urban areas. On the other hand, wealth and educated people are likely to live in urban areas and afford health insurance or out-of-pocket payments for health care [68][69][70]. Media exposure, gender and family norms, women empowerment, female autonomy and community household size may equally be a function of culture, social networks and broader macro-structural underpinnings of society. Media saturation allows better access to information on the importance of antenatal care, facility delivery and postnatal check-ups for mothers and babies. Problematic gender norms and lack of female autonomy are functions of patriarchal systems, which privileges men with power over women and subordinates the status of women [76]. Accordingly, gender division of labour in patriarchal societies is often such that pregnancy and childbirth responsibilities are assigned to women without accompanying social status or access to resources. Thus, making it difficult for them to use available maternal health care services.

The systematic review provides substantial evidence regarding the contribution of contextual factors to maternal healthcare utilisation through the observed ICC measures. Although the study shows mixed results on discriminatory variations in maternal health care between individual and contextual levels, this can be considered in the context of broader literature which suggests that use of maternal healthcare among women are often determined by structural factors beyond their sphere of influence [77]. The relationship between individual-level factors and use of maternal health care services, therefore, could be heavily moderated by community and much more high-level structural factors.

Many attempts have been made to provide theoretical explanations of the relationship between contextual factors and maternal healthcare, albeit with limited success. The lack of reference to social science theory could exacerbate the theoretical deficiencies in maternal health care research. The reasons for this may be the relatively limited number of social science researchers in public health as most of the field is dominated by public health, and epidemiological researchers. Significant theoretical advances have been in social science, especially sociology which could help integrate theory and empirical researcher to better understand underlying mechanisms behind observed complex relationships. The lack of theoretical foundation in public health research is a problem, which has been identified and acknowledged and it is a subject of current debate [71][72].

The strength of this review lies in the fact that it is the first systematic review of literature on multilevel models studying maternal healthcare in SSA. The comprehensiveness of the literature search spanning all relevant databases has given the possibility of reviewing all the relevant literature in selected years to enhance our understanding of the relationships between contextual factors and maternal healthcare in sub-Saharan Africa. The inclusion of the ICC measures helps to discriminate the relative difference in the magnitude of influence between structural conditions and individual characteristics on maternal health care utilisation.

The limitations of the study include the fact that most of the studies selected for this review are cross-sectional and relied on household surveys. There are considerable problems associated with cross-sectional data, and major among them, is failure to provide evidence for causality. Caution should, therefore, be exercised when interpreting the findings of the review and applying the results for policy frameworks. The lack of longitudinal studies in SSA exacerbates this problem. The review has only focussed on quantitative studies and thus underlying reasons behind observable phenomena, which are characteristics of qualitative research studies, have been missed in this review. Additionally, the review only focussed on studies conducted in the English language, so there is potential for studies reported in languages other than English to have been missed. The review restricted itself only to published literature. With the poor publishing culture in SSA, it is probable that there could be a considerable number of studies in the grey literature, which could have added value to this review.

## Conclusion

This study was intended to review the research ecosystem studying the influence of contextual factors on maternal healthcare utilization in sub-Saharan Africa using multilevel techniques. It is foundational work that highlights the contextual factors that are mostly studied, as well as how contextual factor differ from individual factors in influencing maternal healthcare in sub-Saharan Africa. Access and quality of health care services are major determinants of utilization that relate to health systems. Education, poverty, population density, media exposure, gender norms and empowerment at the community level are some of the other important contextual factors. It is important that policy strategies that are aimed at improving maternal health care are focused on these factors.

This study suggests that theoretical pathways of a multilevel nature should always be considered in studies using multilevel techniques to explain how factors operating at different levels interact to influence maternal healthcare utilization. We recommend strong reliance on social science theories and mechanisms to offer substantive explanations of observed relationships between contextual factors and maternal healthcare utilization in SSA.

## Data Availability

All relevant data are within the manuscript and its Supporting Information files

## Author contributions

**Conceptualization:** Simona Simona

**Data curation:** Simona and Casildah Lumamba

**Data extraction and curation:** Simona Simona and Casildah Lumamba.

**Formal analysis:** Simona Simona

**Methodology:** Simona Simona and Million Phiri

`**Software:** Simona Simona and Million Phiri.

**Supervision:** Simona Simona, Casildah Lumamba, Felicitas Moyo, Emily Ng’andu and Million Phiri.

**Validation:** Simona Simona, Felicitas Moyo, Emily Ng’andu and Million Phiri

**Visualization:** Simona Simona

**Writing – original draft:** Simona Simona

**Writing – review & editing:** Simona Simona, Casildah Lumamba, Felicitas Moyo, Emily Ng’andu and Million Phiri

